# Uptake of the Interim Canada Dental Benefit: An investigation of data from the first 18 months of the program

**DOI:** 10.1101/2024.08.19.24312269

**Authors:** Saif Goubran, Vivianne Cruz de Jesus, Anil Menon, Olubukola O. Olatosi, Robert J. Schroth

## Abstract

**Introduction:** In 2022, the Government of Canada introduced the Interim Canada Dental Benefit (CDB) to support Canadian families with children < 12 years of age. This program operated from October 1, 2022, to June 30, 2024, with two application periods. The purpose of this study was to analyze data on applications accepted by the Canada Revenue Agency (CRA) during the first 18 months of the program.

**Methods:** This study used available data sourced from the CRA for applicants as of March 29, 2024, and assessed as of April 5, 2024. Data covered the entirety of the first period (October 1, 2022–June 30, 2023) of the Interim CDB and the first nine months of the second period (July 1, 2023–March 29, 2024). The rate of child participation was calculated using population data from Statistics Canada (2021).

**Results:** Over the first 18 months of the Interim CDB, a total of 410,920 applications were submitted and $403M distributed; $197M for 204,270 applications in period 1 and $175M for 173,160 applications in the first nine months of period 2. A total of 91.8% of applicants had a net family income < $70,000, receiving the maximum benefit amount. The provinces with the highest rate of child participation were Manitoba (77.1/1,000 period 1; 74.9/1,000 period 2), Ontario (82.5/1,000 period 1; 72.2/1,000 period 2), Nova Scotia (73.4/1,000 period 1; 71.1/1,000 period 2), and Saskatchewan (72.3/1,000 period 1; 68.2/1,000 period 2). Overall, there was an increase in the number of applications approved in period 2 compared to period 1.

**Conclusions:** Uptake in the first three quarters of period 2 remained consistent and in many instances, revealed higher rates of applications by parents for the Interim CDB. Analyzing this data will aid in policy recommendation for enhancement of the Canadian Dental Care Program.

## 1 Introduction

While Canada prides itself on its universal health care system, the oral health care needs of many Canadians have seemingly fallen through the cracks. The 2007-2009 Canadian Health Measures Survey revealed that nearly one-third of Canadians lack dental insurance (1). Lower-income families and those without insurance are three to four times more likely to not obtain dental care than higher-income Canadians, with 16.5% declining recommended care due to cost. Furthermore, 56.8% of Canadian children ages 6-11 are affected by dental caries.

Children most at risk of developing caries are those facing barriers to accessing oral health care systems (2). Barriers to accessing care are disproportionately faced by First Nation, Inuit, and Métis children, those living in remote communities, recent refugees and immigrants, and those of low socioeconomic status (3). Children suffering from severe caries face a reduced quality of life due to pain, disturbed sleep, altered growth patterns, low confidence, missed school days, and malnutrition (4). The extent of caries may also warrant dental surgery under general anesthesia, which can be associated with potential complications for young children (5).

In March of 2022, the Canadian government launched a $5.3B program to fund low-income families to cover dental expenses. Parents accessing the Interim Canada Dental Benefit (CDB) received up to a maximum of two payments of up to $650 over the two periods of the program (6-9). The Regular Period 1 of the CDB included eligible children who received dental treatment between October 1, 2022, and June 30, 2023. The Regular Period 2 of the CDB included eligible children who received dental treatment between July 1, 2023, and June 30, 2024.

Only families with annual incomes < $90,000 were eligible to receive the CDB for children <12 years of age (8, 10, 11). Families with private insurance or insurance from an employer-sponsored plan were ineligible to receive the benefit. Applicants must have filed their income tax for the previous year and received the Canada Child Benefit. Applicants receiving Non-Insured Health Benefits or Employment and Income Assistance were also eligible to receive the interim CDB if their dental care expenses for a child were not fully reimbursed. Families with an annual income between $80,000 and $89,999 were eligible to receive $260 for each eligible child. Those with an annual income between $70,000 and $79,999 received $390 for each eligible child. Finally, families with an annual income under $70,000 received $650 for each eligible child. Funds were distributed by the Canada Revenue Agency (CRA).

As the Interim CDB sunset on June 30, 2024, the Canadian Dental Care Plan (CDCP) commenced, providing coverage to a greater range of uninsured Canadians with a family income < $90,000 (8, 12). A national program of such scale presents the opportunity for evaluation and ongoing policy development. The purpose of this study was to analyze data collected by the Government of Canada on applications made and accepted by the Canada Revenue Agency during the entirety of the first period and the first nine months of the second period of the Interim CDB. This study uses available data from the first 18 months of the program sourced from the Canada Revenue Agency for applicants as of March 29, 2024, and assessed as of April 5, 2024.

## 2 Methods

This study analyzed data supplied by the Canada Revenue Agency during the first 18 months of the Interim CDB, up to March 29, 2024, and assessed as of April 5, 2024. Public data was accessed from the Government of Canada Open Data Portal: https://open.canada.ca/data/en/dataset/69035265-2714-4ffa-af3f-fa850209b616 (13, 14). Ethics approval was not required for this quantitative study as it involved exclusively de-identified data publicly accessible from the government of Canada. The Regular Period 1 data include applicants that had dental treatment, applied, and received the Interim CDB between October 1, 2022, to June 30, 2023 (15). The Regular Period 2 data include applicants that had dental treatment, applied, and received the benefit between July 1, 2023, to June 30, 2024 (15).

The Additional Period 1 data include applicants whose children had dental treatment costing exceeded $650 between October 1, 2022, and June 30, 2023 (15). These applicants were eligible to apply and receive an additional $650 within the second pay period, granted they could not apply for the Regular Period 2 of the CDB (15). The Additional Period 2 data include applicants that had children whose dental treatment exceeded $650 between July 1, 2023, and June 30, 2024 (15). These applicants were eligible to apply and receive an additional $650 within the second period, granted they had not applied within the Regular Period 1 of the CDB (15).

Variables of interest include the number of applications submitted to the Canada Revenue Agency, the number of unique applicants, the gender of the approved applicants, the age of the approved applicants, the number of children receiving the benefit, and the total amount of funding received. Such variables were presented as distributed by provinces and territories, age grouping of children, and family net income.

The number of applications and the applicant’s province/territory of residence are obtained from the Canada Dental Benefit file. A unique applicant is an individual. An applicant may apply for more than one child. The residence of the applicant is as of the date of receiving the application. All amounts are rounded to the nearest thousand and are in thousands of dollars, and all counts are rounded to the nearest ten (13, 14). Due to rounding, suppression, and/or double counting for shared custody, the sum of the data may not add to the total (13, 14). Information that has been suppressed for confidentiality purposes is indicated by a “0” (13, 14). Suppressed information also includes valid zeros (13, 14). Individuals whose gender is non-binary are represented in the gender-diverse category (13, 14).

## 3 Results

The number of applications approved by the Interim CDB, the number of unique applicants, and the total amount of funds distributed are reported in Table 1 and Figure 1. During the first 18 months of the Interim CDB, 410,920 applications were submitted, and $403M was distributed. During the Regular Period 1 (October 1, 2022, to June 30, 2023), 204,270 applications made by 188,510 unique applicants for 321,000 children <12 years of age were approved. A total of $197M was distributed by the CRA. During the first nine months of Regular Period 2 (July 1, 2023, to March 29, 2024), there were 173,160 approved applications made by 161,270 unique applicants for 282,130 children <12 years of age. The CRA distributed a total of $175M during this period.

**Table 1.** Number of approved interim CDB applications, unique applicants, and total amount distributed (in $000) by province/territory during Regular Period 1^a^, Regular Period 2^b^, Additional Period 1^c^, and Additional Period 2^d^.

| P/T | Number of Applications | | | | Number of Unique Applicants | | | | Total Amount (\$000) | | | |
| --- | --- | --- | --- | --- | --- | --- | --- | --- | --- | --- | --- | --- |
|  | Regular Period 1 | Regular Period 2 | Additional Period 1 | Additional Period 2 | Regular Period 1 | Regular Period 2 | Additional Period 1 | Additional Period 2 | Regular Period 1 | Regular Period 2 | Additional Period 1 | Additional Period 2 |
| Newfoundland and Labrador | 1,900 | 1,700 | 170 | 210 | 1,720 | 1,590 | 160 | 200 | 1,758 | 1,674 | 143 | 197 |
| Prince Edward Island | 520 | 490 | 40 | 50 | 470 | 450 | 40 | 50 | 470 | 433 | 39 | 44 |
| Nova Scotia | 5,070 | 4,720 | 470 | 610 | 4,590 | 4,330 | 460 | 570 | 4,989 | 4,833 | 430 | 519 |
| New Brunswick | 3,250 | 3,170 | 290 | 420 | 2,920 | 2,880 | 270 | 390 | 3,144 | 3,146 | 253 | 362 |
| Quebec | 37,830 | 30,380 | 1,710 | 2,330 | 34,250 | 27,550 | 1,640 | 2,120 | 33,639 | 28,538 | 1,365 | 1,880 |
| Ontario | 90,940 | 77,170 | 7,130 | 7,770 | 85,030 | 72,830 | 6,920 | 7,280 | 90,695 | 80,069 | 6,544 | 6,873 |
| Manitoba | 9,210 | 8,650 | 830 | 1,300 | 8,100 | 7,760 | 780 | 1,160 | 9,775 | 9,549 | 937 | 1,299 |
| Saskatchewan | 7,800 | 6,990 | 960 | 1,280 | 6,800 | 6,290 | 900 | 1,150 | 8,123 | 7,675 | 1,033 | 1,275 |
| Alberta | 24,140 | 20,250 | 2,060 | 2,480 | 22,340 | 19,010 | 1,990 | 2,300 | 23,862 | 21,107 | 1,931 | 2,287 |
| British Columbia | 23,190 | 19,270 | 1,470 | 1,720 | 21,910 | 18,260 | 1,430 | 1,610 | 20,712 | 17,970 | 1,186 | 1,415 |
| Yukon | 60 | 50 | 0 | 10 | 60 | 50 | 0 | 10 | 51 | 43 | 0 | 7 |
| Northwest Territories | 230 | 190 | 0 | 30 | 200 | 170 | 0 | 30 | 256 | 214 | 0 | 32 |
| Nunavut | 130 | 140 | 50 | 70 | 110 | 120 | 50 | 50 | 133 | 156 | 47 | 66 |
| **Total** | 204,270 | 173,160 | 15,220 | 18,270 | 188,510 | 161,270 | 14,690 | 16,920 | 197,619 | 175,410 | 13,957 | 16,257 |

<sup>a</sup> For services received between October 1, 2022, to June 30, 2023
<sup>b</sup> For services received between July 1, 2023, to March 29, 2024
<sup>c</sup> For applicants who paid more than \$650 for their child's dental services during the Regular Period 1 as of March 29, 2024
<sup>d</sup> For applicants who paid more than \$650 for their child's dental services during the Regular Period 2 as of March 29, 2024

**Figure 1.**
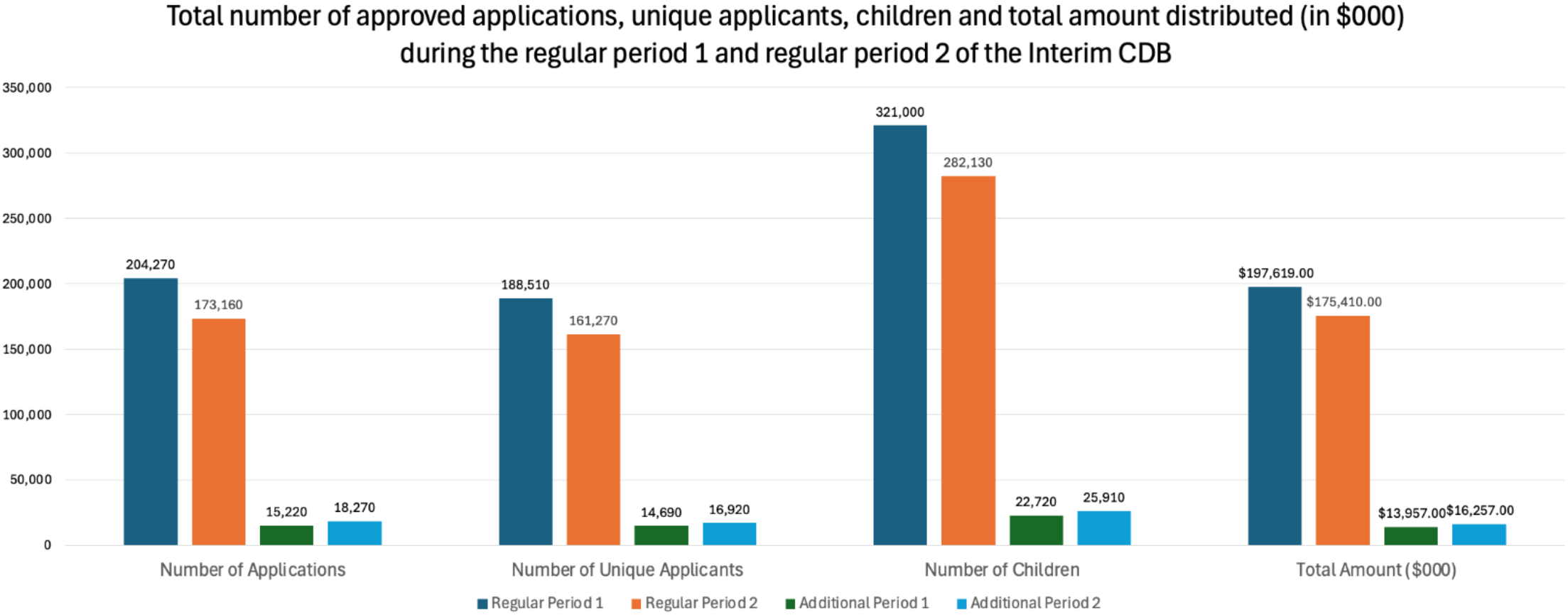
Total number of approved applications, unique applicants, children, and total amount distributed (in $000) during the Regular Period 1^a^, Regular Period 2^b^, Additional Period 1^c^, and Additional Period 2^d^. ^a^ For services received between October 1, 2022, to June 30, 2023 ^b^ For services received between July 1, 2023, to March 29, 2024 ^c^ For applicants who paid more than $650 for their child’s dental services during the Regular Period 1 as of March 29, 2024 ^d^ For applicants who paid more than $650 for their child’s dental services during the Regular Period 2 as of March 29, 2024

During Additional Period 1, 15,220 supplemental applications made by 14,690 unique applicants for 22,720 children were approved (Table 1). A total of $14M was distributed by the CRA. As of March 29, 2024, 18,270 supplemental applications were approved as part of Additional Period 2, made by 16,920 unique applicants for 25,910 children, with the CRA distributing $16M.

As displayed in Table 1, there were a greater number of applicants during the first nine months of the second period of the program when compared to the first period; however, there were a greater number of applicants to the Regular Period 1 of the benefit when compared to the Regular Period 2. Families in Ontario received the greatest proportion of Interim CDB funding in both periods. During the first nine months of the second period of the Interim CDB, over $80M went to Ontario residents (45.6%). Families in the Yukon, Nunavut, and Northwest Territories received the least funding during the second benefit period, totalling $0.413M (0.24%).

Taking into consideration population differences between provinces and territories, the rate of child participation per 1,000 children was calculated using population data from Statistics Canada (2021) for children ages 0-11 and displayed in Table 2 (16). The national rate of child participation was 59.6/1,000 during the first nine months of the Regular Period 2 compared to 67.8/1,000 for the Regular Period 1 (9). The provinces with the highest rate of child participation were Manitoba (77.1/1,000 period 1; 74.9/1,000 period 2), Ontario (82.5/1000 period 1; 72.2/1,000 period 2), Nova Scotia (73.4/1000 period 1; 71.1/1,000 period 2), and Saskatchewan (72.3/1000 period 1; 68.2/1,000 period 2), all having rates of child participation greater than the national rates. Rates of unique applicants to the first nine months of the additional period funding streams (when dental costs exceeded $650) were 4.8/1,000 and 5.5/1,000 for the Additional Period 1 and Additional Period 2, respectively.

**Table 2.** Number of approved Interim CBD children and rate of child participation by province/territory during the Regular Period 1^a^, Regular Period 2^b^, Additional Period 1^c^, and Additional Period 2^d^.

| P/T | Number of Children |  |  |  | Rate of child participation (per 1000) |  |  |  |
| --- | --- | --- | --- | --- | --- | --- | --- | --- |
|  | Regular Period 1 | Regular Period 2 | Additional Period 1 | Additional Period 2 | Regular Period 1 | Regular Period 2 | Additional Period 1 | Additional Period 2 |
| Newfoundland and Labrador | 2,790 | 2,660 | 230 | 310 | 53.3 | 50.8 | 4.4 | 5.9 |
| Prince Edward Island | 780 | 730 | 70 | 80 | 42.6 | 39.9 | 3.8 | 4.4 |
| Nova Scotia | 7,880 | 7,630 | 680 | 820 | 73.4 | 71.1 | 6.3 | 7.6 |
| New Brunswick | 5,050 | 5,040 | 410 | 580 | 58.3 | 58.1 | 4.7 | 6.7 |
| Quebec | 57,400 | 47,930 | 2,360 | 3,100 | 52.2 | 43.6 | 2.1 | 2.8 |
| Ontario | 145,610 | 127,410 | 10,610 | 10,900 | 82.5 | 72.2 | 6.0 | 6.2 |
| Manitoba | 15,530 | 15,080 | 1,490 | 2,020 | 77.1 | 74.9 | 7.4 | 10.0 |
| Saskatchewan | 12,810 | 12,090 | 1,620 | 1,990 | 72.3 | 68.2 | 9.1 | 11.2 |
| Alberta | 39,120 | 34,230 | 3,170 | 3,670 | 60.9 | 53.3 | 4.9 | 5.7 |
| British Columbia | 34,230 | 29,350 | 1,970 | 2,290 | 60.9 | 52.2 | 3.5 | 4.1 |
| Yukon | 90 | 70 | 0 | 10 | 16.5 | 12.8 | 0.0 | 1.8 |
| Northwest Territories | 410 | 340 | 0 | 50 | 59.9 | 49.7 | 0.0 | 7.3 |
| Nunavut | 220 | 240 | 80 | 100 | 22.1 | 24.1 | 8.0 | 10.1 |
| **Total** | 321,000 | 282,130 | 22,720 | 25,910 | 67.8 | 59.6 | 4.8 | 5.5 |

<sup>a</sup> For services received between October 1, 2022, to June 30, 2023
<sup>b</sup> For services received between July 1, 2023, to March 29, 2024
<sup>c</sup> For applicants who paid more than \$650 for their child's dental services during the Regular Period 1
<sup>d</sup> For applicants who paid more than \$650 for their child's dental services during the Regular Period 2

As displayed in Table 3, during the Regular Period 1, 45.6% of unique applicants had a net adjusted family income < $30,000, compared to 54.6% of unique applicants during the first nine months of the Regular Period 2. Over the 18 months of the CDB, 91.8% of applicants had a net family income < $70,000. Overall, 90.8% of children received the maximum benefit amount (i.e., $650) during Period 1 of the program, compared to 92.6% during the first nine months of the second period of the Interim CDB. The proportion of children whose applications for additional funding were approved and received $650 were 92% during the Additional Period 1 and 95% during the Additional Period 2.

**Table 3.**
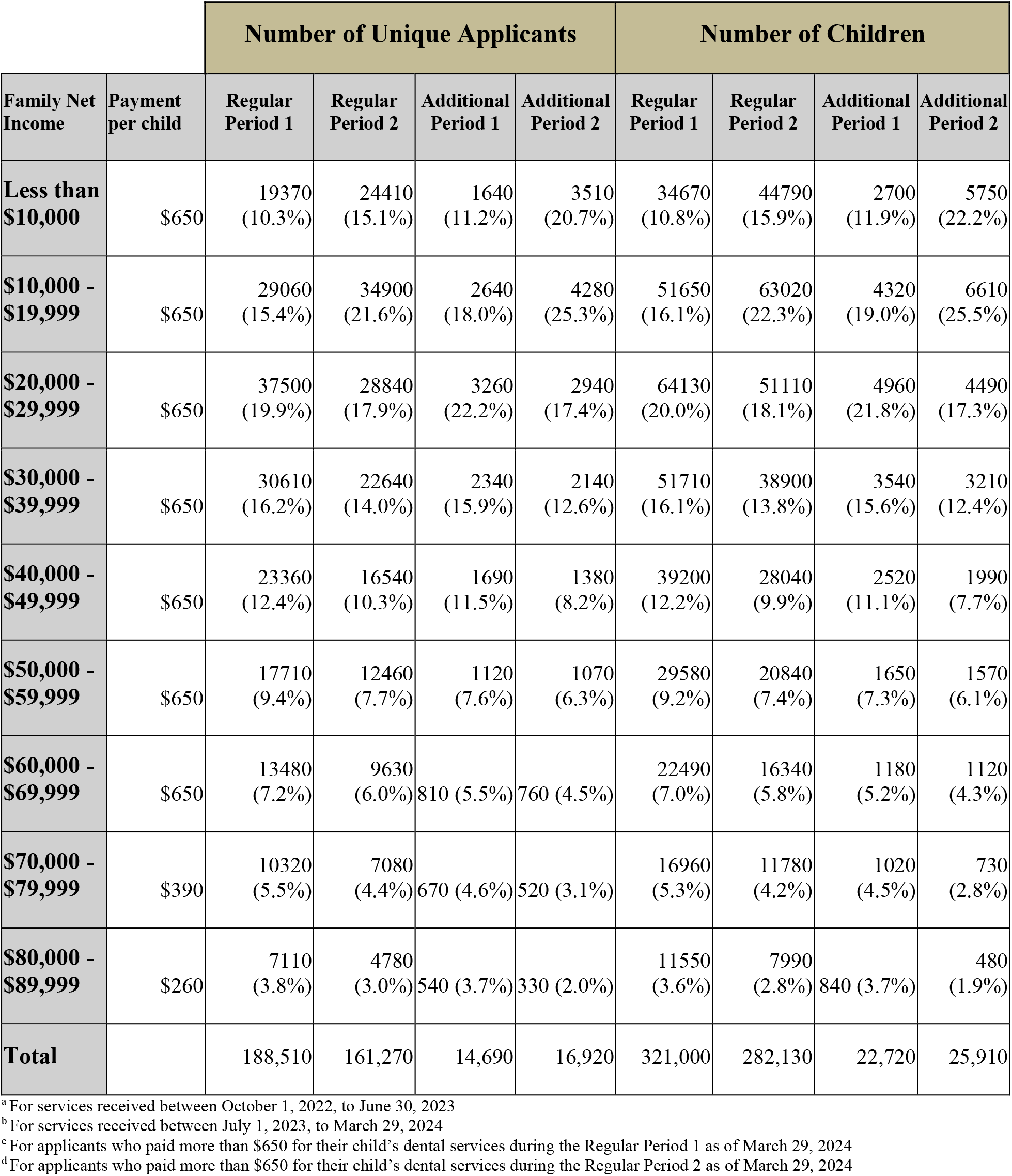
Number of approved Interim CDB applications, unique applicants, children, and total amount (in $000) by adjusted net family income during Regular Period 1^a^, Regular Period 2^b^, Additional Period 1^c^, and Additional Period 2^d^.

Table 4 displays the ages of accepted applicants to the Interim CDB. During Regular Period 1, 83.7% of the applicants were between the ages of 25 and 44 compared to 85% during the first nine months of the second period. Similarly, during the first nine months of additional funding streams, 85% of the applicants were between 25 and 44 (data not shown). The same pattern was true for all provinces and territories.

**Table 4.** Number of approved Interim CDB unique applicants by province and territory, and age group during the Regular Period 1^a^ and Regular Period 2^b^.

| Age group N (%) |  |  |  |  |  |  |  |  |  |  |  |  |  |  |
| --- | --- | --- | --- | --- | --- | --- | --- | --- | --- | --- | --- | --- | --- | --- |
|  | <25 |  | 25-34 |  | 35-44 |  | 45-54 |  | 55-64 |  | 65+ |  | Total<sup>1</sup> |  |
| P/T | RP1 | RP2 | RP1 | RP2 | RP1 | RP2 | RP1 | RP2 | RP1 | RP2 | RP1 | RP2 | RP1 | RP2 |
| Newfoundland and Labrador | 110<br>(6.4) | 80<br>(5.0) | 790<br>(45.9) | 790<br>(49.7) | 680<br>(39.5) | 590<br>(37.1) | 120<br>(7.0) | 110<br>(6.9) | 20<br>(1.2) | 10<br>(0.6) | 0<br>(0.0) | 0<br>(0.0) | 1,720 | 1,590 |
| Prince Edward Island | 10<br>(2.1) | 20<br>(4.4) | 210<br>(44.7) | 200<br>(44.4) | 200<br>(42.6) | 180<br>(40.0) | 60<br>(12.8) | 50<br>(11.1) | 0<br>(0.0) | 0<br>(0.0) | 0<br>(0.0) | 0<br>(0.0) | 470 | 450 |
| Nova Scotia | 250<br>(5.5) | 280<br>(6.5) | 2220<br>(48.4) | 2210<br>(51.0) | 1720<br>(37.5) | 1500<br>(34.6) | 340<br>(7.4) | 300<br>(6.9) | 40<br>(0.9) | 30<br>(0.7) | 20<br>(0.4) | 10<br>(0.2) | 4,590 | 4,330 |
| New Brunswick | 140<br>(4.8) | 170<br>(5.9) | 1410<br>(48.3) | 1390<br>(48.3) | 1110<br>(38.0) | 1070<br>(37.2) | 230<br>(7.9) | 220<br>(7.6) | 10<br>(0.3) | 20<br>(0.7) | 0<br>(0.0) | 0<br>(0.0) | 2,920 | 2,880 |
| Quebec | 590<br>(1.7) | 620<br>(2.3) | 11090<br>(32.4) | 9940<br>(36.1) | 17170<br>(50.1) | 13270<br>(48.2) | 5090<br>(14.9) | 3490<br>(12.7) | 250<br>(0.7) | 190<br>(0.7) | 50<br>(0.2) | 30<br>(0.1) | 34,250 | 27,550 |
| Ontario | 2300<br>(2.7) | 2440<br>(3.4) | 31100<br>(36.6) | 28540<br>(39.2) | 40280<br>(47.4) | 33510<br>(46.0) | 10370<br>(12.2) | 7630<br>(10.5) | 710<br>(0.8) | 500<br>(0.7) | 180<br>(0.2) | 130<br>(0.2) | 85,030 | 72,830 |
| Manitoba | 440<br>(5.4) | 500<br>(6.4) | 3460<br>(42.7) | 3430<br>(44.2) | 3410<br>(42.1) | 3150<br>(40.6) | 680<br>(8.4) | 570<br>(7.3) | 80<br>(1.0) | 100<br>(1.3) | 20<br>(0.3) | 20<br>(0.3) | 8,100 | 7,760 |
| Saskatchewan | 550<br>(1.7) | 550<br>(8.7) | 3040<br>(44.7) | 2920<br>(46.4) | 2580<br>(37.9) | 2280<br>(36.2) | 510<br>(7.5) | 450<br>(7.2) | 90<br>(1.3) | 80<br>(1.3) | 20<br>(0.3) | 10<br>(0.2) | 6,800 | 6,290 |
| Alberta | 720<br>(3.2) | 770<br>(4.1) | 8660<br>(38.8) | 7910<br>(41.6) | 10480<br>(46.9) | 8540<br>(44.9) | 2230<br>(10.0) | 1610<br>(8.5) | 150<br>(0.7) | 120<br>(0.6) | 50<br>(0.2) | 30<br>(0.2) | 22,340 | 19,010 |
| British Columbia | 410<br>(1.9) | 410<br>(2.2) | 6650<br>(30.4) | 6180<br>(33.8) | 11270<br>(51.4) | 9160<br>(50.2) | 3300<br>(15.1) | 2320<br>(12.7) | 190<br>(0.9) | 140<br>(0.8) | 80<br>(0.4) | 40<br>(0.2) | 21,910 | 18,260 |
| Yukon | 0<br>(0.0) | 0<br>(0.0) | 20<br>(33.3) | 20<br>(40.0) | 30<br>(50.0) | 30<br>(60.0) | 0<br>(0.0) | 0<br>(0.0) | 0<br>(0.0) | 0<br>(0.0) | 0<br>(0.0) | 0<br>(0.0) | 60 | 50 |
| Northwest Territories | 10<br>(5.0) | 0<br>(0.0) | 120<br>(60) | 90<br>(52.9) | 60<br>(30.0) | 50<br>(29.4) | 0<br>(0.0) | 0<br>(0.0) | 0<br>(0.0) | 0<br>(0.0) | 0<br>(0.0) | 0<br>(0.0) | 200 | 170 |
| Nunavut | 20<br>(18.2) | 10<br>(8.3) | 60<br>(54.6) | 70<br>(58.3) | 30<br>(27.3) | 30<br>(25.0) | 0<br>(0.0) | 0<br>(0.0) | 0<br>(0.0) | 0<br>(0.0) | 0<br>(0.0) | 0<br>(0.0) | 110 | 120 |
| Total | 5550<br>(2.9) | 5860<br>(3.6) | 68830<br>(36.5) | 63680<br>(39.5) | 89010<br>(47.2) | 73340<br>(45.5) | 22950<br>(12.2) | 16770<br>(10.4) | 1550<br>(0.8) | 1180<br>(0.7) | 440<br>(0.2) | 280<br>(0.2) | 188,510 | 161,270 |

<sup>a</sup> For services received between October 1, 2022, to June 30, 2023
<sup>b</sup> For services received between July 1, 2023, to March 29, 2024
<sup>1</sup> Includes individual applicants with unknown age

A majority of individuals filing applications to the Canada Revenue Agency during the 18 months of the program were female. Over the duration of the program, 362,240 of the 381,390 applicants, or 95.0%, were female. When considering the distribution of funds, $386,699,000 were distributed to female beneficiaries and $15,873,000 to males. Only 36 applicants identified as gender diverse.

Figure 2 reports on the number of children receiving benefits and the total amounts distributed by age during Period 1, the first nine months of Period 2, along with the Additional Periods. As displayed in Figure 2, infants and preschool children accounted for the smallest proportion of recipients of the Interim CDB. During the first nine months of the Regular Period 2, 54.1% of the recipients were less than 6 years of age, receiving $112M. The number of children receiving the benefit and amount received increased with age up to age 4years; this distribution tended to stabilize after the age of 4 years. The highest amount of funding was received by the 7-year-old age group ($19.5M). 11-year-old children received the most Additional Period 1 funding ($4.78M).

**Figure 2.**
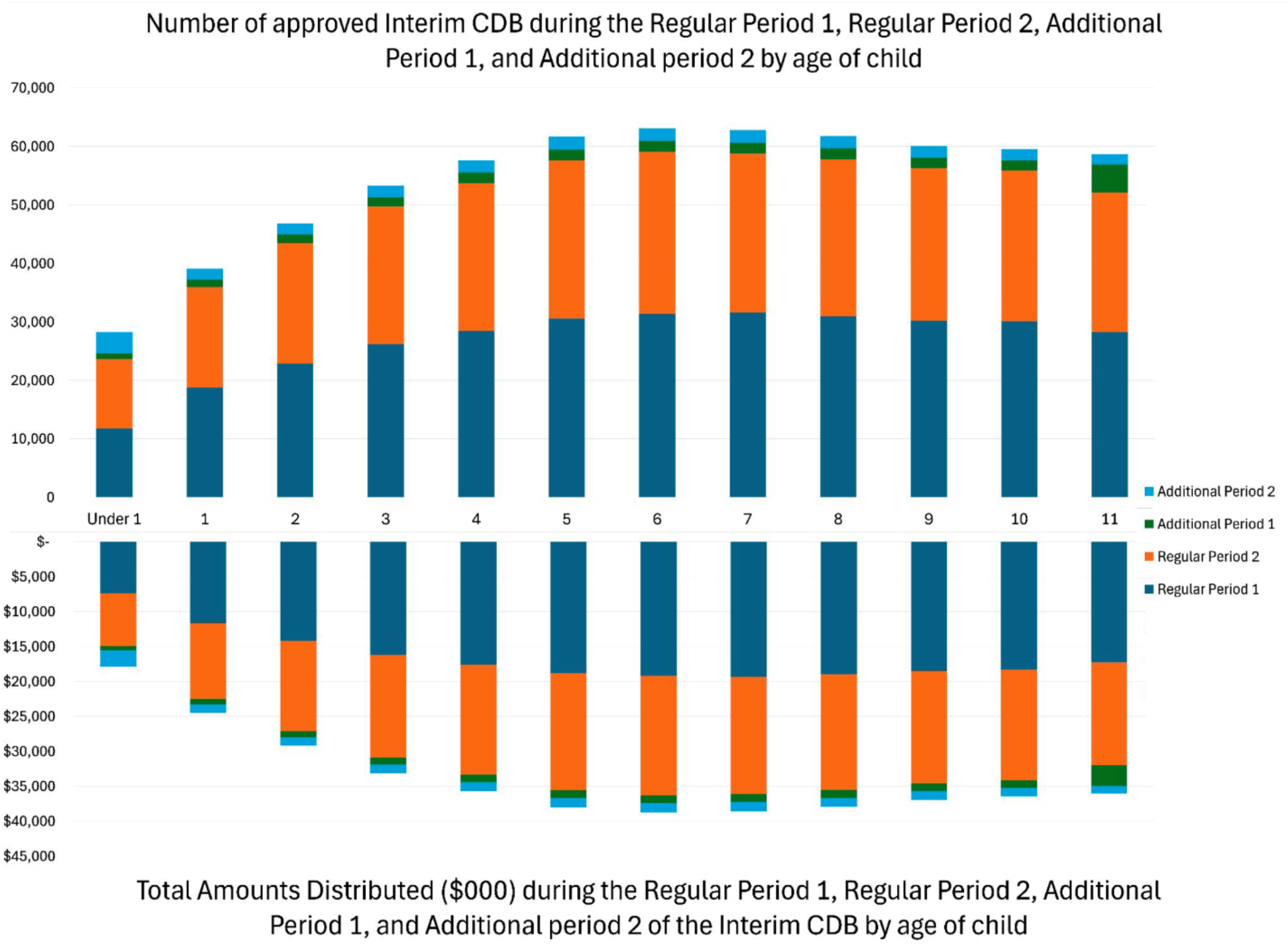
Number of approved Interim CDB children and the total approved amount by age Regular Period 1^a^, Regular Period 2^b^, Additional Period 1^c^, and Additional Period 2^d^. ^a^ For services received between October 1, 2022, to June 30, 2023 ^b^ For services received between July 1, 2023, to March 29, 2024 ^c^ For applicants who paid more than $650 for their child’s dental services during the Regular Period 1 as of March 29, 2024 ^d^ For applicants who paid more than $650 for their child’s dental services during the Regular Period 2 as of March 29, 2024

Table 5 displays the projected totals for the number of applications, number of unique applicants, number of children, and total amount distributed ($000) by June 30, 2024. Projections for the Regular Period 2, Additional Period 1, and Additional Period 2 were calculated by taking the data from the first nine months of the second period (up to March 29, 2024) and projecting it over 12 months (up to June 30, 2024). Using these projections, over 547,000 applications are estimated to be approved between July 1, 2023, and June 30, 2024, made by over 508,000 unique applicants for over 869,000 children <12 years of age. It is projected that the CRA will distribute a total of over $537M.

**Table 5.**
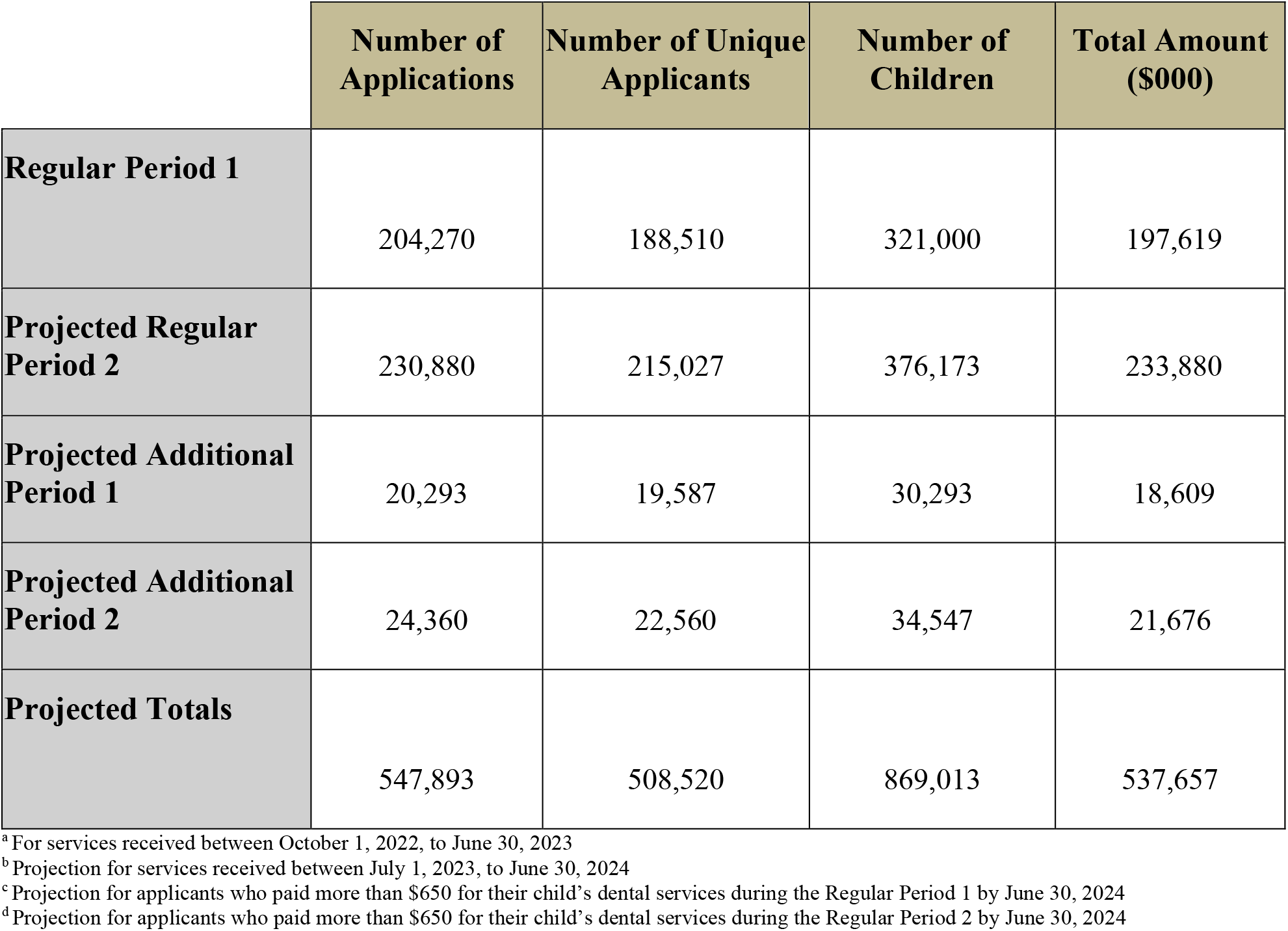
Total number of approved Interim CDB applications, unique applicants, number of children, and total amount distributed (in $000) during the Regular Period 1^a^, and projected totals for Regular Period 2^b^, Additional Period 1^c^, and Additional Period 2^d^ by June 30, 2024.

## 4 Discussion

This study reviewed data from the first 18 months of the Interim CDB program for children < 12 years of age from families with household incomes < $90.000. Overall, our study showed a progressive increase in the number of applications and eligible children benefiting from the Interim CBD program by the end of the program on June 30, 2024. (9). Rates of applications also remained consistent. These findings suggest that there is a proportion of families with financial barriers to accessing oral health care for their children < 12 years of age. The 2007-2009 Canadian Health Measures Survey revealed that nearly one-third of Canadians lack dental insurance (1). Families with no insurance are four times more likely to not obtain dental care compared to higher-income Canadians, with 16.5% declining recommended care due to cost (1). The Canadian Academy of Health Sciences reports that 80% of high-income families have dental insurance, while 50% of low-income families lack dental insurance (3). Those with the fewest barriers to oral health care receive the most support, while those with the greatest need are forced to pay out-of-pocket. A 2020 Ontario study suggests that access to dental insurance increases the proportion of children visiting the dentist (17). Furthermore, the study suggests that access to dental care more significantly impacts the lowest-income groups when compared to the highest-income groups (17). This present study on the Interim CDB corroborates with the findings from this study, showing that children from the lowest-income bracket had the highest uptake of the Interim CDB.

More funds were distributed during the nine months of Regular Period 1 than during the first nine months of Regular Period 2 ($198M compared to $175M). This may be attributed to a lack of awareness that parents were eligible to apply for a second year of the interim CDB for their children, less advertising during Period 2, and even potential dissatisfaction or negative experiences with the program during Period 1. Dental providers may also have been unaware and failed to encourage parents to reapply for the second funding period. It could also be that the growing awareness and media attention regarding the new Canadian Dental Care Plan launching for children < 18 years of age on July 1, 2024, also might have overshadowed Period 2 of the interim CDB. However, when forecasting the full 12 months of Regular Period 2 based on the first nine months, it is estimated that over $233M will be distributed to Canadian families by June 30, 2024. It will be interesting to see if these projections match the final quarter of Period 2 once those data are available from CRA, as families may have misunderstood growing national dentist opposition to the upcoming CDCP as opposition to the interim CDB.

Furthermore, the decrease in Regular Period 2 applicants can be a result of childrens’ dental work exceeding $650, leading families to need to apply for supplemental pay periods during the second period of the benefit as opposed to the Regular Period 2 of the program.

Additional Period 1 applicants include those with dental treatment costing more than $650 during the first period of the benefit, making them eligible to receive an additional $650 within the second pay period, granted they cannot apply for the Regular Period 2 of the CDB. This was the circumstance for 7.01% of children who applied and received insufficient funding to cover their dental costs during the first period of the benefit. As 11-year-old children received the most Additional Period 1 funding, this age group may be experiencing unmet needs more greatly than other age groups, leading to the need for supplemental funding when costs exceeded $650.

Additional Period 2 applicants include those with dental treatments costing more than $650 during the second period of the benefit, making them eligible to receive an additional $650 within the second period, granted they had not applied within the Regular Period 1 of the CDB. Such was the case for 9.18% of children who received the benefit during the second period of the benefit. These individuals may include families that did not apply to the program during the first period. Reasons parents may not have applied for the benefit during the first period can include a lack of awareness due to insufficient advertisement, difficulty understanding the objective of the program due to language barriers, or lack of access to a dental care provider during the time of the first period. Nevertheless, $650 from one period of the benefit was likely inadequate to cover the dental expenses for the child (9).

It has recently been reported that in most Canadian provinces, $650 would likely be able to cover the cost associated with an examination, radiograph, cleaning, fluoride varnish application, and other preventative sealants but likely would not be sufficient to cover the cost of a restorative procedure (9). With children from low-income families being more at risk for caries, $650 may be insufficient to cover the needs of many children (4).

Interestingly, while Regular Period 1 had a greater number of applicants, Regular Period 2 had a greater proportion of applicants with an adjusted family income < $30,000 (45.6% as compared to 54.6%). This could result from an overall decrease in applicants for Regular Period 2 but more awareness of the program amongst low-income families. The increased awareness among low-income families may be attributed to parents being encouraged to apply for the program by community clinics or through word of mouth within the low-income community.

In terms of provincial differences between the number of applications accepted and the amount of funding distributed, the same outcomes are observed between the first and second periods of the benefit (9). The provinces with the highest rate of child participation were Manitoba, Ontario, Nova Scotia, and Saskatchewan. Ontario had the highest participation during Regular Period 1, and Manitoba had the highest participation during Regular Period 2. Results are congruent with reports from Menon et al. 2024, indicating that Manitoba and Saskatchewan may have higher than national rates of participation in the program due to reported poorer access to dental services as compared to other provinces (18). Significantly more parents of children <12 years of age from these two prairie provinces reported access to oral health care challenges than from any other part of the country (18). Furthermore, the national average for the rate of child participation per 1,000 children during Additional Period 1 and Additional Period 2 was 4.8 and 5.5, respectively. Both Manitoba (7.4 and 10.0) and Saskatchewan (9.1 and 11.2) had rates above the national average. The need for additional payments indicates that the treatment of children in these provinces exceeded the $650 distributed, emphasizing the unmet treatment needs of children in these provinces.

Similar to the first period of the benefit, the highest participation was amongst families with a net income less than < $30,000 during the second period of the Interim CDB. Schroth et al. 2024 report that higher-income groups may lack awareness of the benefit, find the program unappealing due to the reduced benefit, or have less need for government support (9).

The territories displayed lower child participation during the first 18 months of the benefit. Such a result may be related to the access to care challenges northern communities face. Schroth et al. (2024)found that, when applying for the Interim CDB, applicants must provide the location of a dental practice they will attend for a scheduled appointment and provide the information of a dental practitioner (9). This requirement may be challenging for individuals living in remote or underserved areas. Given the high proportion of First Nations and Inuit people living in the territories, many children may have federal dental benefits from the Non-Insured Health Benefits Program and may not have seen any benefit in additionally applying for the interim CDB.

Canadians of all ages, particularly children, would benefit from a government-funded program to subsidize the cost of dental care, and the Interim CDB appears to be a great step forward. Figure 2 displays the age groupings of children receiving the Interim CDB. It may come as a welcome surprise that children <1 year of age accounted for 10.3% of children receiving the benefit, as engagement of children this age with the benefit was anticipated to be minimal. Low engagement with this age was anticipated due to a lack of confidence in providers to work with young children as well as a lack of parental awareness of the importance of oral health care for young children.

Findings from this study may be a result of dental offices encouraging parents to apply for the benefit and parents taking ownership to follow through with the application. While the engagement with younger children is impressive, there is still work to be done. Educating parents and oral health care providers on the importance of a first visit prior to the child’s first birthday and the establishment of a dental home can prove to decrease the progression of ECC within communities (19).

This study is not without limitations. As the study uses aggregate data, direct comparisons between the two periods of the benefit are challenging as it is unclear if individuals are new or repeat applicants. Other limitations include an absence of data on the actual oral health care received by children and whether children accessed care beyond an initial exam. However, the applications for the additional payment provide us with some data on children whose initial dental care exceeded $650, suggesting that these children had previous unmet dental needs that have now started to be addressed. Receiving the CDB does not necessarily guarantee that children have all their professional oral health care needs met. Fortunately, the new CDCP will function as insurance and will permit future studies using billing claims data. Overall, this evaluation of the Interim CDB is well aligned with research recommendations from the recently released Canadian National Oral Health Research Strategy 2024-2030 (20).

While programs such as the Interim CDB prove to potentially decrease the financial barrier families face when seeking oral health care, other social determinants decrease the oral health-related quality of life of a child. For instance, families face barriers such as lack of transportation, lack of dental services available in their area, technology barriers, language barriers, poor oral health literacy, and lack of support.

The next directions with the CDCP include increasing program engagement with younger age groups and provinces and territories with low child participation rates. Furthermore, dental offices should continue to encourage low-income families to apply to the program. To effectively improve the oral health-related quality of life of Canadian children, a comprehensive approach must be implemented to address unmet social health determinants.

## 6 Author Contributions

RJS, VCJ, and AM conceptualized the work. SG, VCJ, AM, OOO, and RJS contributed to data analysis and interpretation. All authors contributed to the draft of the manuscript and critically reviewed it. All authors contributed to the article and approved the submitted version.

## 8 Acknowledgments

Robert J Schroth holds Canadian Institutes of Health Research (CIHR) Applied Public Health Chair

## 10 Data Availability Statement

The datasets analyzed for this study can be found in the Government of Canada Open Data Portal: https://open.canada.ca/data/en/dataset/69035265-2714-4ffa-af3f-fa850209b616, through Open Government Licence – Canada.

## Footnotes

1 Includes individual applicants with unknown age

